# Online education and its relation to hearing status among higher-secondary students in Bangladesh: A cross-sectional survey

**DOI:** 10.1101/2024.08.02.24311428

**Authors:** Syeda Tasnim Tabassum Hridi, Mohammad Azmain Iktidar, Arrafi Tamjid, Punam Ghosh, Kazi Sudipta Kabir, Abdullah Al Zaber, Rifat Tasnim Babu, Maliha Mehzabeen, Aysharja Das Gupta, Sreshtha Chowdhury, Simanta Roy

## Abstract

**Background:** Online education gained its popularity in the education system during the COVID-19 pandemic lockdown. The online platform, including social media, was institutionalized globally for the purpose of tutoring to keep the education process ongoing under feasible circumstances. However, the post-pandemic continuation of online education and prolonged usage of electronic devices imposed a greater risk of health issues related to sensory impairment. Our study aimed to determine the impact of online education on students’ hearing status and its associated factors.

**Methods:** A cross-sectional study was conducted among 1030 students of 11th grade and above who were undergoing online education in Dhaka and Chattogram. Data were collected through the online administration of a structured questionnaire containing questions on sociodemographic status, family history of diseases, personal history of comorbidities, information related to screentime exposure, and SSQ-12 (Speech, Spatial, and Qualities of Hearing -12) scale. Descriptive statistics, Pearson’s chi-square test, two independent sample t-tests, and multiple linear regression analysis were employed to obtain the results.

**Result:** The mean SSQ score of the study participants was 7.74±1.37. In bivariate analysis, gender, family income, family history of diseases (e.g., obesity, headache, hearing problem), personal history of diseases (e.g., obesity, insomnia), device type (mobile/tablet, computer), average daily screen time with sound, and break pattern during online learning were significantly (p<0.05 for all) associated with hearing status. In multivariate analysis, being female (coefficient -0.293, p=0.001), using mobile/tablet (coefficient -0.836, p=0.001), and continuous screen use (coefficient -0.348, p=0.003) were significantly associated with poor hearing status.

**Conclusion:** This current study indicates the detrimental effect of online education on the hearing of young students in Bangladesh. Future studies should explore the long-term hearing effects of online education and guide the policy makers towards necessary preventive approaches.

## Introduction

Online education, or virtual learning as an innovative approach, offers the flexibility to access educational resources such as learning materials and lectures with the power of the internet and digital technologies [1]. By breaking down barriers and promoting inclusivity, online education has become a powerful tool in democratizing access to knowledge and empowering learners worldwide [2].

During the COVID-19 pandemic, when the world experienced widespread shutdowns, the adoption of online education skyrocketed to unprecedented levels [3]. This shift affected individuals worldwide, from school-level students to university scholars, who increasingly relied on mobile phones or laptops for virtual learning even after the pandemic [4]. For instance, a cross-sectional study conducted in Bangladesh found that the average length of online education was two to six hours per day [5]. Henceforth, such an extended distance learning period requires prolonged use of headphones and speakers, which could ultimately lead to hearing issues like noise-induced hearing loss [4].

For online education, students often relied on headphones or speakers, varying sound quality, and diverse learning environments with sub-optimal acoustic conditions [6]. The factors mentioned above potentially contributed to prolonged exposure to sound at unsuitable levels, leading to an increased risk of hearing-related issues [7]. A nationwide survey in South Korea found that one in five adolescents using earphones for at least 80 minutes daily showed signs of hearing loss [8]. Another cross-sectional study among adults in Andra Pradesh, India, revealed that using headphones for multiple purposes, such as leisure, education, service, music, and gaming, resulted in subclinical hearing loss [9]. In addition, using earphones in a noisy environment contributed to over four times higher hearing impairment among adolescents [8] and led to tinnitus, characterized by ringing or buzzing in the middle ear cavity [10].

Since gadgets for online education and leisure activities have witnessed exponential growth, the risk of auditory health problems has become a pressing concern. Although age, heredity, gender, and ethnicity have been identified as factors contributing to noise-induced hearing loss[11], there remains a dearth of comprehensive data regarding the specific impact of online education on hearing, which often involves continuous and regular exposure to noises. Therefore, this study aims to address this gap by investigating the effects of online education on the hearing status of Bangladeshi students.

## Methods

### Study design, sites, and participants

A cross-sectional survey was conducted between 25 May 2022 and 31 July 2023 to assess the impact of online education on the hearing status of the students studying in 11th grade and above. Preselected educational institutes, including tuition homes or academies located in Dhaka and Chattogram, were chosen to obtain data that considered the accessibility of students from diverse educational backgrounds. For the students who were 18 years and above, the informed consent was directly obtained from them. However, for participants below 18 years, the informed consent was obtained from their parents. The consent form contained all the detailed information concerning the study and the probable outcome of participation. The study did not involve the requirement of any sensitive information.

### Sampling and selection criteria

A total of 1030 students were enrolled in the study through convenient sampling. The aforementioned sampling strategy was applied considering the limitation of resources. Even though the sampling technique lacks randomization, the large sample size (n=1030) helped to overcome this limitation by reducing the error margin, leading to more precise results. The following inclusion criteria were considered before selecting the study participants: (1) students who provided voluntary consent for participation, (2) students who were citizens of Bangladesh, and (3) students who received online education during the period of COVID-19 lockdown and after. We excluded students under the following criteria: (1) foreign students studying in Bangladesh, (2) students with previous histories of hearing impairment, and (3) students who were engaged with online education before COVID-19 lockdown.

### Data Collection Techniques

A structured questionnaire was prepared for online administration through Google Forms. The students were asked to answer the questions after providing their consent electronically for participation. The questionnaire included information on sociodemographic status (age, gender, educational qualification, income), family history of diseases, personal history of comorbidities, information related to screentime exposure (daily hours spent on electronic devices for study and entertainment, type of device used, breaks taken during use, duration of online education in months), and information regarding their hearing status. Mandatory items were highlighted with a red asterisk, and the relevant non-response option was also incorporated. Respondents could review their answers through the back button and change their responses if necessary. The survey was never displayed a second time once the user had filled it in to prevent duplicate entries. Of the 1050 eligible participants who agreed to participate, 1030 participants completed the entire questionnaire (completion rate: 98.09%); incomplete questionnaires were excluded from the analysis.

### Hearing assessment

The primary outcome of the study was the hearing status of the students which was measured via Speech, Spatial and Qualities of Hearing Scale questionnaire (SSQ-12) [12]. The scale included 12 items under 4 sub-scales: speech scale (items 1, 2, 3, 4, 5) spatial scale (items 6, 7, 8) and qualities of hearing scale (items 9. 10, 11, 12). For each item in each sub-scale there is a score that runs from 0 to 10. The students needed to identify with the score that resembles their own experiences to the described situation in the item and select that score. The score provided a quantitative measure of hearing disability in each domain. The lower the hearing experience, the lower the average score.

### Statistical Analysis

The obtained data were analyzed using Stata (v.17.0) statistical software. The results were tabulated in frequencies and corresponding percentages for the categorical variables. However, mean with standard deviations were reported for continuous variables. Associations between dependent and independent variables were assessed using Pearson’s chi square test and t-test. A linear regression model was used to determine the factors associated with hearing problems. Any statistical values were deemed significant at p<0.05.

### Ethics

The study proposal was reviewed and approved by the institutional review board of North South University (approval no: 2022/OR-NSU/IRB/0403). The objective, risks, and benefits of the research were well explained to all the study participants before participation. The anonymity of the participants was maintained, and they were allowed to withdraw from the study at any time of the study period. Wherever feasible, the 1964 Declaration of Helsinki and later modifications and comparable ethical standards were followed. Data collection was voluntary, and no incentives were offered to participants. Data were only accessible to the authors and were not disclosed anywhere. All the reporting was done according to the Checklist for Reporting Results of Internet E-Surveys (CHERRIES) guidelines[13].

## Results

A total of 1030 students participated in the study. Their background information is presented in Table 1. The mean age of the participants was 18.62±3.04 years, and the majority of them were female (60.58%). Two-thirds of the students were studying in 12th grade, and one-third of the students had their family’s monthly income ranging between thirty thousand to sixty thousand bdt ($300-600). The majority of the students had a family history of comorbidities (82.72%), with only 8.54% having a family history of hearing problems. Eye problems were reported as a major concern among the students (68.74%). Most of the students (60.78%) were continuing online education for more than 12 months, and mobile/tablets were reported as the most used electronic device for both education (94.56%) and entertainment (92.62%). The majority of the students had a screen time of 2-6 hours (66.67%) for online education and less than 2 hours (43.69%) for entertainment.

**Table 1:** Background information of the study participants.

| Variables | Frequency | Percentage |
| --- | --- | --- |
| **Age** |  |  |
| <18 years | 359 | 46.44 |
| ≥18 years | 414 | 53.56 |
| **Gender** |  |  |
| Male | 406 | 39.42 |
| Female | 624 | 60.58 |
| **Education** |  |  |
| 11<sup>th</sup> grade | 24 | 2.33 |
| 12<sup>th</sup> grade | 664 | 66.8 |
| Undergraduate | 258 | 25.05 |
| Graduate and post-graduate | 84 | 8.16 |
| **Family income (in bdt)** |  |  |
| <10,000 | 96 | 9.32 |
| 10,000 to 30,000 | 312 | 30.29 |
| 30,000 to 60,000 | 384 | 37.28 |
| 60,000 to 1,00,000 | 174 | 16.89 |
| >1,00,000 | 64 | 6.21 |
| **Family history of diseases** |  |  |
| Comorbidity | 852 | 82.72 |
| Obesity | 234 | 22.72 |
| Headache | 538 | 52.23 |
| Eye Problem | 708 | 68.74 |
| Insomnia | 346 | 33.59 |
| Hearing Problem | 88 | 8.54 |
| **Personal history of diseases** |  |  |
| Obesity | 134 | 13.01 |
| Headache | 448 | 43.5 |
| Eye Problem | 708 | 68.74 |
| Insomnia | 152 | 14.76 |
| **Headphone use** |  |  |
| No | 368 | 35.73 |
| Yes | 662 | 64.27 |
| **Online education-related use** |  |  |
| **Duration (months)** |  |  |
| 1 to 3 months | 28 | 2.72 |
| 3 to 6 months | 74 | 7.18 |
| 6 to 12 months | 302 | 29.32 |
| >12 months | 626 | 60.78 |
| **Used mobile/ tablet** |  |  |
| No | 56 | 5.44 |
| Yes | 974 | 94.56 |
| **Used computer** |  |  |
| No | 666 | 64.66 |
| Yes | 364 | 35.34 |
| **Used TV** |  |  |
| No | 1,008 | 97.86 |
| Yes | 22 | 2.14 |
| **Screentime with sound/day** |  |  |
| <2 hours | 130 | 12.67 |
| 2-6 hours | 684 | 66.67 |
| 6-12 hours | 174 | 16.96 |
| >12 hours | 38 | 3.7 |
| **Break pattern** |  |  |
| Use with enough break<br>(15 min break after every 2 hours of use) | 282 | 27.38 |
| Use with a small break<br>(10 min break after every 2 hours of use) | 476 | 46.21 |
| Continuous use (4 hours without break) | 272 | 26.41 |
| **Online entertainment related use** |  |  |
| **Used mobile/ tablet** |  |  |
| No | 76 | 7.38 |
| Yes | 954 | 92.62 |
| **Used computer** |  |  |
| No | 754 | 73.2 |
| Yes | 276 | 26.8 |
| **Used TV** |  |  |
| No | 748 | 72.62 |
| Yes | 282 | 27.38 |
| **Screentime with sound/day** |  |  |
| <2 hours | 450 | 43.69 |
| 2-6 hours | 432 | 41.94 |
| 6-12 hours | 118 | 11.46 |
| >12 hours | 30 | 2.91 |
| **Break pattern** |  |  |
| Use with enough break<br>(15 min break after every 2 hours of use) | 454 | 44.08 |
| Use with a small break<br>(10 min break after every 2 hours of use) | 376 | 36.5 |
| Continuous use (4 hours without break) | 200 | 19.42 |

However, shorter break time was noted in the majority (46.21%) of the students during online education, and most of the participants (64.27%) used headphones while using their digital devices.

Table 2 presents the bivariate relationship between the participant and device use characteristics and mean SSQ score among the study population. Male participants had a significantly higher SSQ score compared to the female counterparts (7.93 vs 7.61, p<0.001). Participants with family history of comorbidity (p<0.001), obesity (p<0.001), headache (p<0.001), insomnia (p<0.001), hearing problem (p=0.021), and personal history of obesity(p=0.006), psychological problem (p<0.001), insomnia (p<0.001) had significantly lower SSQ score. Screen time for online education (p=0.014), mobile (p<0.001), and computer (p<0.001) usage for online education, break pattern during online education (p=0.013), and entertainment (p=0.006) were also significantly associated with SSQ score.

**Table 2:** Bivariate relationship between SSQ score and participant characteristics.

| Variable Name | SSQ Score |  | p-value |
| --- | --- | --- | --- |
|  | Mean | SD |  |
| **Age (years)** |  |  | 0.3592 |
| <18 | 7.77 | 1.29 |  |
| $\geq 18$ | 7.68 | 1.41 | |
| **Gender** |  |  | <0.001 |
| Male | 7.93 | 1.21 |  |
| Female | 7.61 | 1.45 |  |
| **Education** |  |  | 0.2054 |
| 11<sup>th</sup> grade | 7.44 | 1.31 |  |
| 12<sup>th</sup> grade | 7.72 | 1.37 |  |
| Undergraduate | 7.73 | 1.34 |  |
| Graduate and post-graduate | 8.01 | 1.47 |  |
| **Income (in bdt)** |  |  | <0.001 |
| <10,000 | 7.61 | 1.68 |  |
| 10,000 to 30,000 | 7.60 | 1.40 |  |
| 30,000 to 60,000 | 7.77 | 1.38 |  |
| 60,000 to 1,00,000 | 7.92 | 1.11 |  |
| >1,00,000 | 7.95 | 1.22 |  |
| **Family history of diseases** |  |  |  |
| **Comorbidity** |  |  | <0.001 |
| No | 8.09 | 1.11 |  |
| Yes | 7.67 | 1.41 |  |
| **Obesity** |  |  | <0.001 |
| No | 7.83 | 1.35 |  |
| Yes | 7.42 | 1.40 |  |
| **Headache** |  |  | <0.001 |
| No | 8.00 | 1.22 |  |
| Yes | 7.50 | 1.45 |  |
| **Insomnia** |  |  | **<0.001** |
| No | 7.87 | 1.34 |  |
| Yes | 7.47 | 1.53 |  |
| **Hearing Problem** |  |  | **0.02** |
| No | 7.77 | 1.35 |  |
| Yes | 7.42 | 1.53 |  |
| **Personal history of diseases** |  |  |  |
| **Comorbidity** |  |  | 0.55 |
| Yes | 7.72 | 1.42 |  |
| No | 7.78 | 1.20 |  |
| **Obesity** |  |  | **0.006** |
| No | 7.78 | 1.34 |  |
| Yes | 7.44 | 1.54 |  |
| **Headache** |  |  | 0.07 |
| No | 7.81 | 1.28 |  |
| Yes | 7.65 | 1.48 |  |
| **Insomnia** |  |  | **<0.001** |
| No | 7.82 | 1.33 |  |
| Yes | 7.25 | 1.50 |  |
| **Headphone use** |  |  | 0.28 |
| No | 7.80 | 1.41 |  |
| Yes | 7.70 | 1.35 |  |
| **Online Education-Related Variables** |  |  |  |
| **Duration (months)** |  |  | 0.05 |
| 1 to 3 months | 6.57 | 1.78 |  |
| 3 to 6 months | 7.81 | 1.37 |  |
| 6 to 12 months | 7.65 | 1.42 |  |
| >12 months | 7.82 | 1.30 |  |
| **Used mobile/tablet** |  |  | **<0.001** |
| No | 8.34 | 1.18 |  |
| Yes | 7.70 | 1.37 |  |
| **Used computer** |  |  | **<0.001** |
| No | 7.62 | 1.40 |  |
| Yes | 7.96 | 1.30 |  |
| **Used TV** |  |  | **0.57** |
| No | 7.74 | 1.37 |  |
| Yes | 7.58 | 1.25 |  |
| **Screentime with sound/day** |  |  | **<0.001** |
| <2 hours | 7.51 | 1.54 |  |
| 2 to 6 hours | 7.84 | 1.23 |  |
| 6 to 12 hours | 7.55 | 1.65 |  |
| >12 hours | 7.70 | 1.53 |  |
| **Break pattern** |  |  | **0.007** |
| Use with enough break<br>(15 min break after every 2 hours of use) | 7.90 | 1.35 |  |
| Use with a small break<br>(10 min break after every 2 hours of use) | 7.75 | 1.28 |  |
| Continuous use (4 hours without break) | 7.56 | 1.52 |  |
| **Online Entertainment Related Variables** |  |  |  |
| **Used mobile/tablet** |  |  | **0.65** |
| No | 7.67 | 1.40 |  |
| Yes | 7.74 | 1.37 |  |
| **Used computer** |  |  | **0.26** |
| No | 7.71 | 1.40 |  |
| Yes | 7.82 | 1.29 |  |
| **Used TV** |  |  | **0.07** |
| No | 7.69 | 1.43 |  |
| Yes | 7.86 | 1.20 |  |
| **Screentime with sound/day** |  |  | **0.14** |
| <2 hours | 7.77 | 1.39 |  |
| 2 to 6 hours | 7.73 | 1.31 |  |
| 6 to 12 hours | 7.69 | 1.44 |  |
| >12 hours | 7.63 | 1.70 |  |
| **Break pattern** |  |  | 0.89 |
| Use with enough break<br>(15 min break after every 2 hours of use) | 7.89 | 1.38 |  |
| Use with a small break<br>(10 min break after every 2 hours of use) | 7.58 | 1.35 |  |
| Continuous use (4 hours without break) | 7.71 | 1.35 |  |

Table 3 presents the results from multiple linear regression to investigate the factors associated with the SSQ score. The results showed that, after adjusting for all other variables, gender was significantly related to the SSQ score. The SSQ score was 0.293 units lower in females than males (β = -0.293, 95% CI: -0.461 to -0.124, p = 0.001). Participants without a family history of comorbidities had 0.277 units higher SSQ scores than those with such a history (β = 0.277, 95% CI: 0.055 to 0.499, p = 0.015). Similarly, individuals without a family history of obesity exhibited higher SSQ scores of 0.310 units than those with such a history (β = 0.310, 95% CI: 0.110 to 0.510, p = 0.003). In terms of duration of use, compared to 1 to 3 months, engaging in online education for 3 to 6 months was associated with a 1.276-unit increase in SSQ score (β = 1.276, 95% CI: 0.700 to 1.853, p < 0.001). Similarly, durations of 6 to 12 months and over 12 months were associated with SSQ score increases of 1.065 units (β = 1.065, 95% CI: 0.551 to 1.579, p < 0.001) and 1.295 units (β = 1.295, 95% CI: 0.791 to 1.799, p < 0.001) respectively. Conversely, using mobile or tablet devices for online education corresponded to lower SSQ scores of 0.836 units compared to not using such devices (β = -0.836, 95% CI: -1.228 to -0.445, p < 0.001). In terms of break patterns, compared to using devices with sufficient breaks (15 minutes after every 2 hours of use), a continuous usage pattern for 4 hours without breaks was associated with a decrease in SSQ score by 0.348 units (β = -0.348, 95% CI: -0.578 to -0.117, p = 0.003). For entertainment purposes, using mobile or tablet devices resulted in higher SSQ scores of 0.477 units higher than not using them (β = 0.477, 95% CI: 0.137 to 0.816, p = 0.006), whereas using a TV resulted in SSQ scores that were 0.185 units higher than not using it (β = 0.185, 95% CI: 0.002 to 0.370, p = 0.049).

**Table 3:** Multiple linear regression results for the predictors of SSQ score.

| Variables | Coefficient | p-value | Lower bound of 95% CI | Upper bound of 95% CI |
| --- | --- | --- | --- | --- |
| **Gender** |  |  |  |  |
| Male | Ref |  |  |  |
| Female | -0.293 | **0.001** | -0.461 | -0.124 |
| **Family history of comorbidities** |  |  |  |  |
| Yes | Ref |  |  |  |
| No | 0.277 | **0.015** | 0.055 | 0.499 |
| **Family history of obesity** |  |  |  |  |
| Yes | Ref |  |  |  |
| No | 0.31 | **0.003** | 0.11 | 0.51 |
| **Online Education-Related Variables** |  |  |  |  |
| **Duration (months)** |  |  |  |  |
| 1 to 3 | Ref |  |  |  |
| 3 to 6 | 1.276 | **<0.001** | 0.700 | 1.853 |
| 6 to 12 | 1.065 | **<0.001** | 0.551 | 1.579 |
| >12 | 1.295 | **<0.001** | 0.791 | 1.799 |
| **Using mobile or tablet** |  |  |  |  |
| No | Ref |  |  |  |
| Yes | -0.836 | **<0.001** | -1.228 | -0.445 |
| **Break Pattern** |  |  |  |  |
| Use with enough break<br>(15 min break after every 2 hours of use) | Ref |  |  |  |
| Use with a small break<br>(10 min break after every 2 hours of use) | -0.151 | 0.134 | -0.349 | 0.047 |
| Continuous use (4 hours without break) | -0.348 | **0.003** | -0.578 | -0.117 |
| **Online Entertainment Related Variables** |  |  |  |  |
| **Using mobile or tablet** |  |  |  |  |
| No | Ref |  |  |  |
| Yes | 0.477 | **0.006** | 0.137 | 0.816 |
| **Using TV** |  |  |  |  |
| No | Ref |  |  |  |
| Yes | 0.185 | 0.049 | 0.002 | 0.370 |
| **Break pattern** |  |  |  |  |
| Use with enough break<br>(15 min break after every 2 hours of use) | Ref |  |  |  |
| Use with a small break<br>(10 min break after every 2 hours of use) | -0.264 | **0.005** | -0.451 | -0.078 |
| Continuous use (4 hours without break) | -0.018 | 0.881 | -0.256 | 0.219 |

p-values in bold=statistically significant; CI=Confidence interval

## Discussion

The unprecedented rise in the adoption of online educational platforms has unveiled many concerns, encompassing the potential consequences on the auditory well-being of students. The study was conducted using a self-reported questionnaire showing that online education’s rapid rise causes hearing impairments. During the pandemic, digital learning became a daily requirement, resulting in a significant increase in the usage of digital devices among students. The world is currently witnessing a surge in online education, which has increased digital device usage. The current research, the first in the region, examines the possible factors associated with hearing among students continuing online education.

Gender differences showed significant variation, with females showing more susceptibility to poor hearing. Similar findings were also reported by another study in which women reported more subjective hearing problems attributable to headphone usage than men. [14] The underlying reason could be genetic, hormonal [15], or environmental. Also, better hearing was reported among students from high socio-economic status, which might be attributable to better awareness, ambient acoustic environment, and use of devices of better quality.

Individuals with a family history of comorbidities were more prone to experiencing hearing issues, aligning with prior research indicating an increased association of hearing loss with conditions such as obesity,[16, 17], and insomnia[18] at an individual level, rather than solely in first-degree relatives. This suggests that a positive family history or genetic predisposition consistently correlates with a higher likelihood of hearing complications.

Interestingly, respondents with shorter experiences of online education had worse hearing complications than those with longer duration. This finding mirrors previous studies that identified a relationship between the duration of device use and hearing impairment.[19–21] According to the studies, the explanation could be increasing participants’ awareness of health issues and their physiological adaptation over time. They might have followed precautions upon experiencing hearing issues. However, longitudinal studies are required to have a definite answer to this question.

Furthermore, this study showed that using portable hand-held devices such as mobile and tablets caused a greater impact on respondents’ hearing impairments than using larger-sized devices such as TVs. The distance between the user and devices could be the reason behind this [22]. Despite lacking data on individual noise exposure, it can be said that hand-held devices run close to the users and are often used with hearing equipment (e.g., headphones), which can affect hearing more adversely than other mediums.

Continuous exposure to sound can irritate the hair cells in the inner ear and cause sustained damage to hearing. Online education involves prolonged and continuous exposure to sounds, which can impair hearing. This current study found a declining trend in hearing among participants who were continuously using gadgets for online education. This finding is consistent with another study that found prolonged headphone use disrupted the high-frequency threshold and significantly impaired balance in adolescents who spent more than eight hours a day in front of a screen [23]. Therefore, ensuring sufficient breaks in using the gadgets can be helpful as a preventive strategy.

The findings were notably different when it came to entertainment purposes, with smartphone and tablet use linked to less hearing damage than when it came to educational use. This could be explained by the fact that respondents were expected to be more focused on listening to online classes, possibly with the highest volume for educational purposes. For entertainment purposes, the use could be more intermittent and relaxed, and perhaps optimal volume could be used as needed, which might explain why the same type of devices caused two opposite impacts on the hearing of respondents, depending on the individual’s purpose. Furthermore, optimum distance from devices with external audio devices plays a crucial role in auditory health. This study showed that TV played a less harmful role in hearing health when used for entertainment because of the distance from respondents and the delivery of efficient sound frequency.

A limitation of this study was the use of a non-probability sampling approach due to limited resources, which may limit the generalizability of findings. However, we included more than double the required samples from diverse backgrounds to keep the error minimum. Secondly, specialist assessment of hearing is the most standard method of assessing hearing, which couldn’t be done. However, we used a validated tool (sensitivity 85.7% and specificity 86.1%) to assess hearing, which was used in many previous studies. Lastly, causality cannot be established due to the cross-sectional nature of the study design. Despite limitations, this is the first study to provide valuable insights about hearing among students continuing online education in Bangladesh with a large sample size. The novel character of our study is further highlighted by the lack of a thorough body of previous research, which calls for more investigation to validate and build upon our results.

## Conclusion

This current study revealed that online education was related to hearing impairment among students due to prolonged and continuous exposure to noise. Given the inescapable rise in screen usage among students, it is worth considering increasing efforts to educate students about the appropriate use of electronic devices in order to minimize the adverse effects.

## Data Availability

Data cannot be shared publicly because of participants' confidentiality and institutional restriction. Data are available from the North South Institutional Data Access / Ethics Committee for researchers who meet the criteria for access to confidential data. However, the data can be provided to the editorial board members upon request.

## Acknowledgments

We are grateful to Zerine Mahjabin Chowdhury for her assistance in the research. We would also like to mention Dr. Azaz Bin Sharif for his generosity and support.

## Author Contributions

Conceptualization: STTH. Data Curation: PG. Formal Analysis: MAI. Methodology: SR. Project Administration: STTH, SC. Supervision: SR, MAI. Visualization: RTB. Manuscript Original Draft Preparation: KSK, AT, MM, ADG. Manuscript Review & Editing: KSK, MAI, SR.

## Notes

### Competing Interest Statement

The authors have declared no competing interest.

### Funding Statement

The author(s) received no specific funding for this work.

### Author Declarations

The study proposal was reviewed and approved by the Institutional Review Board of North South University (approval no: 2022/OR-NSU/IRB/0403).

